# Differential Performance of CoronaCHEK SARS-CoV-2 Lateral Flow Antibody Assay by Geographic Origin of Samples

**DOI:** 10.1101/2021.04.12.21255284

**Authors:** Owen R. Baker, M. Kate Grabowski, Ronald M. Galiwango, Aminah Nalumansi, Jennifer Serwanga, William Clarke, Yu-Hsiang Hsieh, Richard E. Rothman, Reinaldo E. Fernandez, David Serwadda, Joseph Kagaayi, Tom Lutalo, Steven J. Reynolds, Pontiano Kaleebu, Thomas C. Quinn, Oliver Laeyendecker

## Abstract

**Background:** We assessed the performance of CoronaCHEK lateral flow assay on samples from Uganda and Baltimore to determine the impact of geographic origin on assay performance.

**Methods:** Serum samples from SARS-CoV-2 PCR+ individuals (Uganda: 78 samples from 78 individuals and Baltimore: 266 samples from 38 individuals) and from pre-pandemic individuals (Uganda 1077 and Baltimore 532) were evaluated. Prevalence ratios (PR) were calculated to identify factors associated with a false-positive test.

**Results:** After first positive PCR in Ugandan samples the sensitivity was: 45% (95% CI 24,68) at 0-7 days; 79% (95%CI 64,91) 8-14 days; and 76% (95%CI 50,93) >15 days. In samples from Baltimore, sensitivity was: 39% (95% CI 30, 49) 0-7 days; 86% (95% CI 79,92) 8-14 days; and 100% (95% CI 89,100) 15 days post positive PCR. The specificity of 96.5% (95% CI 97.5,95.2) in Ugandan samples was significantly lower than samples from Baltimore 99.3% (95% CI 98.1,99.8), p<0.01. In Ugandan samples, individuals with a false positive result were more likely to be male (PR 2.04, 95% CI 1.03,3.69) or individuals who had a fever more than a month prior to sample acquisition (PR 2.87, 95% CI 1.12,7.35).

**Conclusions:** Sensitivity of the CoronaCHEK was similar in samples from Uganda and Baltimore. The specificity was significantly lower in Ugandan samples than in Baltimore samples. False positive results in Ugandan samples appear to correlate with a recent history of a febrile illness, potentially indicative of a cross-reactive immune response in individuals from East Africa.

## INTRODUCTION

Severe acute respiratory syndrome coronavirus 2 (SARS-CoV-2) infection causes coronavirus disease 2019 (COVID-19) (1), which has been detected on all continents and continues to be a public health emergency globally (2). Critical to public health efforts to combat the pandemic are accurate serologic assays to differentiate exposed from unexposed individuals (3). Many studies investigate the performance of these assays on samples from Asia (4), Western Europe (5), and the United States (6). However, little information is available on the performance of these assays in an African setting, though initial studies provide evidence of potential problems (7), particularly among febrile patients infected by other infectious pathogens (8).

Serologic assays used for the detection of antibodies to different viral infections can vary in performance based on the origin of the samples being tested, as has been seen in HIV (9), HCV (10), and HSV-2 (11). It is thought that these differences in specificity result from host genetics of the source population and the frequency and distribution of the infectious agents exposed to the population (12). We sought to compare the performance of the CoronaCHEK Lateral Flow Assay (LFA) on samples from Uganda and the United States to assess the impact of geographic origin on the performance of this assay. Samples from known SARS-CoV-2 infected individuals with known duration of infection and pre-pandemic samples were tested to evaluate the sensitivity and specificity of the assay and to identify factors associated with a false positive result.

## METHODS

### Ethics statement

The use of samples from Baltimore was approved by The Johns Hopkins University School of Medicine Institutional Review Board (IRB00247886, IRB00250798, and IRB00091667). The use of samples from Uganda was approved by the Uganda Virus Research Institute’s Research Ethics Committee (GC/127/20/04/773, GC/127/13/01/16), Western Institutional Review Board, protocol 200313317 and the Uganda National Council for Science and Technology (HS637ES). The parent studies were conducted according to the ethical standards of the Helsinki Declaration of the World Medical Association, where all subjects provided written informed consent. All samples were de-identified prior to testing.

### Sample sets

To assess sensitivity, samples from subjects known to be SARS-CoV-2 PCR+ from Uganda and the United States with known duration from first PCR+ date were evaluated. Samples from 78 PCR+ individuals at different time intervals were identified at the Uganda Virus Research Institute in Entebbe, and Makerere University in Kampala, Uganda. None of the Ugandan individuals were hospitalized and all had mild disease. Samples (n=266) from the United States were from 38 hospitalized COVID-19 patients, attending the Johns Hopkins Hospital in Baltimore, Maryland in the United States (13).

To assess the specificity of the assay, pre-pandemic samples were tested. This included 1077 stored samples from the Rakai Community Cohort Study, collected between 2011 and 2013 (14). The Ugandan samples included 543 individuals who reported having been febrile within the month prior to sample acquisition and 534 individuals who did not report a febrile illness, matched by age and gender. The 532 pre-pandemic samples from the US were remnant CBC samples collected from Johns Hopkins Hospital Emergency Department (JHH ED) patients collected between December 2015 and January 2016 (15).

### Laboratory Testing and Statistical Analysis

All samples were analyzed with the CoronaCHEK LFA (Hangzhou Biotest Biotech Co Ltd) according to the manufacturer’s protocol. Sensitivity by duration of infection and specificity among pre-pandemic samples were assessed for the presence of either IgM or IgG bands for any reactivity. Statistical analysis was performed with STATA 14.2 (Statacorp College Station, Texas, USA), and 95% confidence intervals (95% CI) for sensitivity and specificity were calculated with the Clopper-Pearson exact method. Bivariate Poisson regression models were used to calculate prevalence ratios (PR) for factors associated with a false-positive test among pre-pandemic samples.

## RESULTS

There were significant differences in the performance for the CoronaCHEK LFA between samples from Uganda and Baltimore (**Table 1**). When comparing any reactivity (IgM or IgG) there was no significant difference in reactivity by duration of infection. Though 100% of samples from Baltimore were seropositive by 14 days after their first time point, this was not the case for the Ugandan samples. Specificity, when considering any reactive band as a false positive result, was significantly lower in Ugandan samples at 96.9% (CI 95.2, 97.5) than in those from Baltimore, 99.3% (CI 98.1, 99.8), p<0.01. When limited to Ugandan samples collected from individuals with no reported febrile illness in the month prior to sample collection (n=500), the specificity was still significantly lower 96.8% (CI 95.0,98.1) than in those samples from Baltimore, p<0.05.

**Table 1.** Sensitivity and Specificity of CoronaCHEK Lateral Flow Point of Care Assay for the Detection of IgM and IgG Antibodies to SARS-CoV-2

| **Sensitivity** | **Performance** |  |  |
| --- | --- | --- | --- |
|  | IgM % (95% CI) | IgG % (95% CI) | IgM or IgG % (95% CI) |
| Uganda |  |  |  |
| ≤ 7 days (n=22) | 41% (21 - 64) | 41% (21 - 64) | 45% (24 - 68) |
| >7 to 14 days (n=39) | 74% (58 - 87) | 49% (32 - 87) | 79% (64 - 91) |
| >14 – 28 days (n=17) | 41% (18 - 67) | 65% (38 - 86) | 76% (50 - 93) |
| Baltimore |  |  |  |
| ≤ 7 days (n=102) | 34% (25 – 44) | 21% (13 – 30) | 39% (30 – 49) |
| >7 to 14 days (n=132) | 82% (74 – 88) | 75% (67 – 82) | 86% (79 – 92) |
| >14 – 28 days (n=32) | 100% (89 – 100) | 100% (89 – 100) | 100% (89 – 100) |
| **Specificity** |  |  |  |
| Uganda (n=1077) | 96.9% (95.7 - 97.9) | 99.4% (98.7 - 99.7) | 96.5% (95.2 - 97.5) |
| Baltimore (n=532) | 99.3% (98.1 – 99.8) | 100% (99.3 - 100) | 99.3% (98.1 - 99.8) |

There were four and 38 false positive results in Baltimore pre-pandemic samples and Ugandan samples, respectively. All four from Baltimore were all faint IgM bands while 82% (31/38) of the false positive samples from Uganda had only reactive IgM bands. Of the seven pre-pandemic Ugandan samples that were IgG reactive, two were also reactive for IgM. Ugandan samples were significantly more likely to misclassify if they came from men (PR 2.04, 95% CI 1.03, 3.69, p=0.04) or the individual had reported fever more than a month prior to sample collection (PR 2.87, 95% CI 1.12, 7.35, p=0.028). There was a trend to test positive if they had reported pneumonia-like symptoms (PR 2.34, 95% CI 0.98, 5.59, p=0.056). Other factors not associated with a false positive result included age, community type, and HIV status (**Table 2**). There were too few misclassified samples from Baltimore to assess factors associated with misclassification within this population.

**Table 2.** Factors associated with a false positive SARS-CoV-2 antibody response in samples from Uganda.

| Defining Category | Outcome: SAR-CoV-2 Antibody Positive |  |
| --- | --- | --- |
|  | % (n/N) | PR (95% CI) |
| ***Categorical variables*** |  |  |
| Sex |  |  |
| *Female* | 2.7% (20/737) | Ref. |
| *Male* | 5.3% (18/340) | 2.06 (1.03, 3.69) |
| Age |  |  |
| 18-24 | 3.1% (10/327) | Ref. |
| 25-34 | 4.3% (19/439) | 1.42 (0.66, 3.04) |
| 35-44 | 2.7% (7/260) | 0.88 (0.34, 2.31) |
| 45-54 | 3.9% (2/61) | 1.28 (0.28, 5.85) |
| Community Type |  |  |
| *Agrarian* | 3.2% (14/436) | Ref. |
| *Fishing* | 5.1% (19/372) | 1.59 (0.80, 3.17) |
| *Trading* | 1.9% (5/269) | 0.58 (0.21, 1.61) |
| Pregnancy (no males in analysis) |  |  |
| *Not pregnant* | 2.5% (8/318) | Ref. |
| *Pregnant* | 2.9% (12/419) | 1.14 (0.47, 2.78) |
| Fever < 1 mo |  |  |
| *No* | 3.2% (17/534) | Ref. |
| *Yes* | 3.9% (21/543) | 1.21 (0.64, 2.30) |
| Fever > 1 mo |  |  |
| *No* | 3.2% (33/1,023) | Ref. |
| *Yes* | 9.3% (5/54) | 2.87 (1.12, 7.35) |
| Cough |  |  |
| *No* | 3.3% (27/825) | Ref. |
| *Yes* | 4.4% (11/252) | 1.33 (0.66, 2.69) |
| Pneumonia |  |  |
| *No* | 3.2% (32/997) | Ref. |
| *Yes* | 7.5% (6/80) | 2.34 (0.98, 5.59) |
| HIV Status |  |  |
| *Negative* | 3.4% (21/618) | Ref. |
| *Positive* | 3.7% (17/459) | 1.09 (0.58, 2.07) |

## DISCUSSION

This study demonstrates differential performance of the CoronaCHEK LFA on samples collected from Uganda compared to those collected from Baltimore. Though sensitivity for both IgG and IgM in samples from Baltimore was 100% by 14 days after the subjects first PCR+ date, unlike samples from Uganda, this difference was not significantly different. Specificity was significantly lower in the Ugandan pre-pandemic samples compared to those from Baltimore, though this difference was all associated with the IgM band. False positive results in Ugandan samples were higher among men and those who had reported a febrile episode more than a month prior to sample acquisition. Of the false positive results detected, the vast majority were IgM reactivity.

These results demonstrate that the performance characteristics of serological assays for SARS-CoV-2 antibody detection cannot be extrapolated to different populations without adequate validation studies. This study supports the need for validation studies on SARS-CoV-2 serologic assays in Africa, an area where little data exists (16). Though a lower specificity was found in Ugandan samples than those from Baltimore, the specificity of 96.5% was much greater than the 85% found for the Euroimmun IgG S1 ELISA in pre-pandemic samples from Benin (8). As shown in the study by Mboumba Bouassa (7), our study demonstrated that the main cause for false positive results was a reactive IgM test. If one ignores the presence of an IgM band, the specificity of the CoronaCHEK increased to 99.4% (95% CI 98.7, 99.7) for Ugandan samples and 100% (95% CI 99.3, 100) in Baltimore samples, with no loss of sensitivity at 14 days post first positive PCR for SARS-CoV-2.

There are a number of limitations of our study. First, the samples from Uganda of SARS-CoV-2 infected patients were limited, with only six samples within the first week post first PCR positive test and no serial samples for a given individual. Additionally, these samples from known infected Ugandan individuals had limited symptoms, while the Baltimore samples from known SARS-CoV-2 positive individuals were all hospitalized subjects. The pre-pandemic samples from Baltimore were not matched to those from Uganda based on symptomology, though historically, individuals attending the ED in the United States have a high prevalence of fever and viral infections (17). Samples from the JHH ED do have a high burden of chronic viral infections, as demonstrated by a seroprevalence of 6%, 12% and 50% for HIV, HCV and HSV-2 respectively (18).

In summary, the geographical origin of the samples appeared to impact the performance of the CoronaCHEK LFA. IgM reactivity was the main cause for the false positive results. Given that IgM responses generally appear a couple days before IgG, it may be useful not to measure IgM at all in serological studies given the improvement in specificity. Further evaluations of serologic assays are needed to find appropriate tools for sero-surveillance in an African setting.

## Data Availability

Data is available upon request

## Acknowledgements

We acknowledge all of the participants who contributed specimens to this study and the study staff without whom this study would not have been possible.

## Notes

Funding: Support was provided by the Division of Intramural Research, National Institute of Allergy and Infectious Diseases (NIAID), National Institutes of Health (NIH), as well as by extramural support from NIAID UM1-AI068613 for supporting R.E.F.; and NIH Center of Excellence in Influenza Research and Surveillance HHSN272201400007C to R.E.R.

### Competing Interest Statement

The authors have declared no competing interest.

### Funding Statement

Support was provided by the Division of Intramural Research, National Institute of Allergy and Infectious Diseases (NIAID), National Institutes of Health (NIH), as well as by extramural support from NIAID UM1-AI068613 for supporting R.E.F.; and NIH Center of Excellence in Influenza Research and Surveillance HHSN272201400007C to R.E.R.

### Author Declarations

The Johns Hopkins University School of Medicine Institutional Review Board (IRB00247886, IRB00250798, and IRB00091667); Uganda Virus Research Institute Research Ethics Committee (GC/127/20/04/773, GC/127/13/01/16); Western Institutional Review Board (200313317); Uganda National Council for Science and Technology (HS637ES).

